# High PrEP uptake and objective longitudinal adherence among HIV-exposed women with personal or partner plans for pregnancy in rural Uganda

**DOI:** 10.1101/2022.08.10.22278611

**Authors:** Lynn T Matthews, Esther C Atukunda, Moran Owembabazi, Kato Paul Kalyebera, Christina Psaros, Pooja Chitneni, Craig W Hendrix, Mark A Marzinke, Peter L Anderson, Oluwaseyi O Isehunwa, Kathleen E Wirth, Kara Bennett, Winnie Muyindike, David R Bangsberg, Jessica E Haberer, Jeanne M Marrazzo, Mwebesa Bosco Bwana

## Abstract

**Background:** In Uganda, fertility rates and adult HIV prevalence are high, and many women conceive with partners living with HIV. Oral pre-exposure prophylaxis (PrEP) reduces HIV acquisition risk for women and, therefore, infants. We conducted a longitudinal cohort study in Uganda to evaluate oral PrEP uptake and adherence as part of HIV prevention in the context of reproductive goals for women (i.e., safer conception care).

**Methods:** We enrolled HIV-negative women with personal or partner plans for pregnancy with a partner living, or thought to be living, with HIV (2017-2020) to evaluate the impact of the Healthy Families intervention on PrEP use. Quarterly study visits through 9 months included HIV and pregnancy testing, and safer conception counseling. PrEP was provided to women in electronic pillboxes as the primary adherence measure (categorized as “high” with pillbox openings on ≥80% of days). Enrollment questionnaires assessed factors associated with PrEP use. Plasma tenofovir (TFV) and intraerythrocytic TFV-diphosphate (TFV-DP) concentrations were determined at each visit for women who acquired HIV during follow-up and a randomly selected subset of those who did not. Women who became pregnant were initially exited from the cohort by design; from April 2019, women with incident pregnancy remained in the study with quarterly follow-up until pregnancy outcome. Primary outcomes included (1) PrEP uptake (proportion of enrolled women who initiated PrEP) and (2) PrEP adherence (proportion of days with electronic pillbox openings during the first 3 months following PrEP initiation). We used univariable and multivariable-adjusted linear regression to evaluate baseline predictors of mean adherence over 3 months. We also assessed mean monthly adherence over 9 months of follow-up and during pregnancy.

**Results:** We enrolled 131 women with a mean age of 28.7 years (95% CI: 27.8-29.5). Ninety-seven (74%) reported a partner with HIV and 79 (60%) reported condomless sex at last encounter. Most women (N=118; 90%) initiated PrEP. Mean electronic adherence during the 3 months following initiation was 87% (95% CI: 83%, 90%); most (85%) had adherence >80%. No covariates were associated with 3-month pill-taking behavior. Plasma TFV concentrations were ≥40ng/mL among 66%, 56%, and 45% at months 3, 6, and 9, respectively. TFV-DP concentrations were ≥600 fmol/punch among 47%, 41%, and 45% of women at months 3, 6, and 9. We observed 53 pregnancies among 131 women with 848 person-months of follow-up (annualized incidence 75% [95% CI: 57%, 98%]) and one HIV-seroconversion in a non-pregnant woman accessing PrEP. Mean pillcap adherence for PrEP users with pregnancy follow-up (N=17) was 98% (95% CI: 97%, 99%).

**Conclusions:** Women in Uganda with PrEP indications and planning for pregnancy chose to use PrEP. By electronic pillcap, most were able to sustain high adherence to daily oral PrEP prior to and during pregnancy. Differences in adherence measures highlight challenges with adherence assessment and serial measures suggest 41-66% of women took sufficient periconception PrEP to prevent HIV. These data suggest that women planning for and with pregnancy should be prioritized for PrEP implementation, particularly in settings with high fertility rates and generalized HIV epidemics.

## Introduction

Despite declining overall HIV prevalence in Uganda, median antenatal HIV prevalence remains high at 6%-7% (1). Uganda has one of the highest total fertility rates in the world at 4.7 children per woman (2), and while services to prevent perinatal transmission are robust for pregnant women with HIV, HIV prevention prior to a desired pregnancy is rarely addressed. However, at least 30-50% of men living with HIV in Uganda desire children (3–7) and nearly half have a stable, HIV-negative partner (8). Women risk condomless sex to meet important personal and sociocultural goals to have children (3, 7, 9–11). In 2019, over 20,000 women of reproductive age and 5,700 children were newly diagnosed with HIV in Uganda, with perinatal transmission accounting for most infections (12). Integrating HIV prevention into reproductive health programs presents an opportunity to reduce HIV incidence among women and infants in settings where fertility rates and HIV prevalence are high. Indeed, prevention in this context is relevant for many women across sub-Saharan Africa where the average fertility rate is 4.6 (13, 14).

Effective HIV prevention strategies are available to women who want to conceive with a partner living with HIV including delaying condomless sex until the partner achieves viral load suppression by taking antiretroviral therapy (ART), treating sexually transmitted infections (STI) in both partners, limiting condomless sex to peak fertility, and semen processing (15–17). However, in settings where gender power imbalances make it challenging for a woman to insist that her partner participate in strategies to reduce sexual HIV transmission and where many men are not aware of their status, pre-exposure prophylaxis (PrEP) is an important safer conception strategy. Data suggest that tenofovir disoproxil fumarate (TDF) and emtricitabine (FTC) are safe to use during early pregnancy, and with high adherence, PrEP can nearly eliminate HIV acquisition risks (18–20).

The World Health Organization and the Ugandan Ministry of Health (MoH) recommend PrEP as a preventive approach for HIV-negative individuals at high-risk of acquiring HIV, including women partnered with someone living with HIV (21, 22). PrEP implementation in periconception and antenatal settings has been low (23), and data on PrEP uptake and adherence among women planning for pregnancy are limited (24, 25). The few observational studies and clinical trials exploring PrEP use and adherence among specific populations of women, including adolescent girls, young women, and those in HIV-serodifferent couples, in East and Southern Africa document varying acceptability and uptake of PrEP ranging from 8% among adolescent girls to 100% among sexually-active women without stated plans to become pregnant (26–30). PrEP adherence studies of women at risk for HIV during pregnancy also observe variable adherence rates based on pharmacy pick-up as well as drug levels, ranging from 22%- 62% at 3 months (28, 31, 32). Understanding how women initiate and adhere to PrEP as periconception risk reduction is an important step towards developing comprehensive HIV prevention care for women of reproductive age (33, 34).

Based on formative work and informed by a conceptual framework for periconception HIV-exposure behavior (35), we developed a counseling support intervention, Healthy Families-PrEP, for HIV-negative women of reproductive age in Uganda with personal or partner plans for a pregnancy in the next year. Healthy Families-PrEP leverages individual- and couple-level reproductive goals to promote uptake and use of HIV prevention strategies, including TDF/FTC PrEP. We followed women to evaluate uptake of and use of PrEP during periconception and pregnancy periods.

## Methods

### Study Design and Population

Women enrolled between June 2017 and January 2019. Recruitment took place in rural, southwestern Uganda from a safer conception pilot program located within the Mbarara Regional Referral Hospital (36), HIV counselling and testing sites in the district, and via referrals from local healthcare providers. Women also approached the program after hearing about it via flyers, community testing events, and informational radio spots.

Eligible women were aged 18-40 years, tested negative for HIV (rapid test), not currently pregnant (urine b-HCG testing), likely to be fertile (based on reproductive health history) (37), and reported personal or partner desire to have a child in the next year (38–41). Additionally, an eligible woman either knew her pregnancy partner was living with HIV or felt she was at risk for acquiring HIV(42). All enrolled women provided informed consent and felt able to attend study visits for the duration of the study.

### Study Procedures

Participants received a package of HIV prevention or safer conception counselling for women who want to conceive a child while exposed to HIV, Healthy-Families PrEP (Figure 1). Study visits occurred quarterly and included HIV testing, pregnancy testing, PrEP adherence counseling, and safer conception counselling sessions.

**Figure 1.**
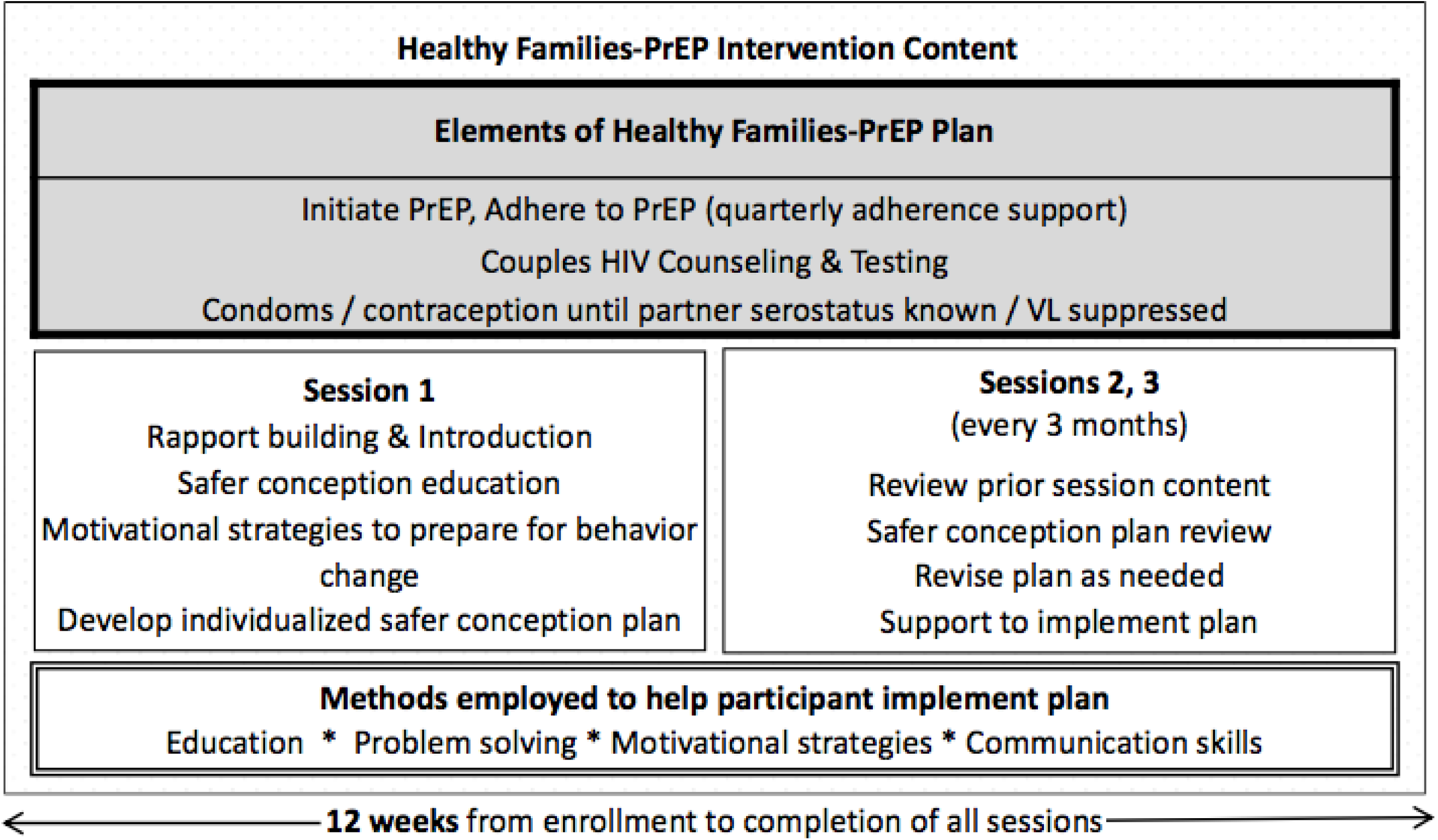
Intervention Content From the Healthy Families-PrEP Program

The Healthy Families-PrEP intervention was informed by a socio-ecological framework to understand women’s PrEP use (43, 44), and our periconception risk behavior conceptual framework (35). Healthy Families-PrEP leveraged individual- and couple-level reproductive goals to promote uptake and use of HIV prevention strategies through education, problem-solving, communication-skills training, and adherence support. Trained counselors worked with participants to develop and implement a safer conception plan. For women who did not know their partner’s HIV status, this plan included how to encourage their partners to test and disclose their status. For those with partners living with HIV, counselling included how to encourage partners to initiate ART, to delay condomless sex until partner achieved either ART coverage for 6 months or HIV viral load suppression, and to limit sex without condoms to peak fertility. Oral combination TDF (300mg) /FTC (200mg) as Truvada was offered as PrEP and adherence counseling was included at quarterly visits based on methods developed by Dr. Psaros and colleagues (45).

Support for each of these strategies was available at the clinic study site including couples-based counseling and testing, condoms, ART, and contraception. Ovulation prediction kits were offered by the study. In addition, participants were counselled regarding opportunities for sperm washing, donor sperm, and adoption as alternatives available in other parts of Uganda.

At baseline, women completed a questionnaire that included measures on socio-demographic, health status, reproductive history, HIV knowledge, safer conception behaviors, and other constructs expected to impact PrEP uptake and adherence based on our periconception risk reduction conceptual framework (35). Women were eligible to initiate TDF/FTC as PrEP at any time during study follow-up. Through April 2019, women who became pregnant during follow-up had final study evaluations at the time of first positive pregnancy test. They were exited from the study with referrals for antenatal and routine PrEP care. Based on ethical concerns about pregnant women not wanting to access PrEP through the public sector, the protocol was updated. From March 2019, women who became pregnant during study follow-up remained in the study and were followed every 3 months until a pregnancy outcome occurred. For women with incident pregnancies after March 2019, the final visit was conducted after the pregnancy outcome. At all final visits, women were referred for PrEP care in the public sector if desired. Women who tested positive for HIV during study follow up completed exit activities at the time of first positive HIV test and were referred to appropriate HIV follow-up care.

All participants completed quarterly urine pregnancy tests (beta-HCG), rapid fourth generation HIV1/2 screening (and confirmation as indicated) per Ugandan standard of care, and syndromic screening for STIs. A subset of women was screened for asymptomatic STI including Chlamydia trachomatis, Neisseria gonorrhoeae, and Trichomonas vaginalis via GeneXpert testing and syphilis via treponemal and non-treponemal antibody testing at baseline and 6 months follow-up. Participants who screened or tested positive for STIs received treatment per local guidelines; STI prevalence and incidence findings are reported elsewhere (46). Blood was drawn at baseline for creatinine and hepatitis B assessment to ensure no contraindications to PrEP use. Creatinine concentrations were assessed during quarterly follow-up.

Women with abnormal renal function (serum Creatinine >89 µmol/L and/or GFR <60 mL/min estimated using the Cockcroft-Gault equation) or active hepatitis B infection (HBV surface antigen positive) were subsequently instructed to discontinue PrEP.

### Measures

As part of questionnaires, data were collected on socio-demographic characteristics (age, education, and socio-economic status), reproductive health history (number of prior pregnancies, live births and living children), sexual behavior (number of sexual partners, condom use and sexual encounters), and HIV disclosure within the pregnancy partnership. Questionnaire responses were collected to construct the following scores: knowledge of HIV (47) and of safer conception behaviors (48), HIV risk perception (42), PrEP optimism (adapted) (49), parenthood motivation (50), reproductive autonomy (51), sexual relationship power (52), partner communication (53), functional social support (54), and depression (55, 56). Summary scores were derived using cited methods (see Appendix).

### Assessment of PrEP Uptake and Adherence

To measure daily pill-taking behavior, women were provided with an electronic pillbox (Wisepill Technologies, South Africa) that stored PrEP tablets and recorded when the device was opened, providing a reliable, objective assessment of day-to-day adherence behavior (57). We assessed two primary outcomes: (1) uptake of PrEP defined as the proportion of enrolled women who ever initiated PrEP and (2) objective adherence to PrEP as measured by the proportion of days with pillbox opening during the first 3 months of active PrEP follow-up among PrEP initiators. Adherence was defined as the number of days with a time-stamped record of a device opening divided by the number of days the participant was in active PrEP follow-up (defined as PrEP initiation through to the earlier of reported PrEP discontinuation or study exit) and capped at one opening per day. We chose to focus on 3-month adherence given high pregnancy incidence resulting in meaningful changes in the population over time. We also report on mean monthly adherence over time and the proportion of PrEP initiators with monthly adherence ≥80%, categorized as “high” adherence (58, 59). These data are also described for women with pregnancy as secondary analyses.

Due to the potential for adherence misclassification (e.g., curiosity openings of the pillbox without dosing, device non-use), biomedical assessments of PrEP adherence were also determined for a subset of participants via plasma TFV and dried blood spot (DBS) intraerythrocytic TFV-DP concentrations. All women who ever-accessed PrEP had blood samples drawn at quarterly visits. Plasma TFV was processed for women who acquired HIV during study follow-up and a random subset of women who did not acquire HIV. TFV-DP was processed for all collected samples. TFV and TFV-DP levels were quantified using via liquid chromatographic-tandem mass spectrometric (LC-MS/MS) analysis using previously described methods (64, 69–71).

Plasma TFV levels reflect dosing in the last week: concentrations >40ng/mL indicate dosing in the last 24 hours (categorized as “high” adherence), between 10 and 40 ng/mL indicates dosing in the last 3 days (categorized as “moderate” adherence), and between >0.31 and 10 ng/mL indicates dosing in the last week (categorized as “low” adherence) (20, 60). TFV concentrations below 0.31 ng/mL are below the limits of detection. TFV-DP reflects average dosing in the last 6-8 weeks; >600 fmol/punch indicates approximately ≥4 doses per week (categorized as high adherence), 450-600 fmol/punch indicates about 3-4 doses per week (categorized as moderate adherence), and between 31.3 and <450 indicates <3 doses per week (categorized as low adherence) (57, 61–63). TFV-DP concentrations below < 31.3 fmol/punch are below limits of detection.

### Statistical analysis

We conducted univariable and multivariable-adjusted analyses to assess predictors of mean PrEP adherence during the first 3 months following PrEP initiation. Baseline covariates were selected based on our periconception HIV risk conceptual framework (35) and included age, education, number of live births, depression, parenthood motivation (as related to social control), sexual relationship power scale, reproductive autonomy (decision-making subscale), and perceived HIV risk. For each baseline predictor (relative to the adherence outcome), we separately constructed multivariable-adjusted models using a change-in-estimate approach (64). Specifically, relative to a fully adjusted model (i.e., all known and/or hypothesized confounding factors included), we removed, one by one, each factor and recorded the estimated log relative risk (RR) for the predictor-outcome association of interest. If the removal of the factor changed the log RR by ≥10%, it was retained in the final multivariable model. Given the large number of results only the final adjusted RR (and 95% CI) for each predictor of interest is presented. A complete description of each multivariable-adjusted model is included in the appendix.

For secondary analyses of adherence over 6 and 9 months of follow-up, we fit intercept-only modified Poisson regression models with generalized estimating equations stratified by month to estimate mean monthly adherence and 95% CI. Overall adherence over 6 and 9 months was calculated by taking an inverse variance weighted mean of monthly estimates to account for variable amounts of person-time contributing to each monthly estimate. We performed Spearman correlation analysis to assess the relationship between plasma TFV and whole blood drug TFV-DP concentrations and electronic pillcap adherence at each of the 3-month follow-up visits (through 9 months) where data was available. Lastly, we performed exploratory analyses to assess adherence to PrEP before and after date of first positive pregnancy test until the reported date of pregnancy outcome. All statistical analyses were conducted using SAS software version 9.4 (SAS Institute, Cary, NC).

### Ethics Statement

Ethics approvals were secured from the Partners HealthCare Institutional Review Board (IRB), Mbarara University of Science and Technology Research Ethics Committee, and University of Alabama at Birmingham IRB. Regulatory approvals were also secured from Uganda’s Office of the President and the National Council of Science and Technology.

## Results

Of the 916 women who were screened, 131 (14%) met study criteria and enrolled. The study was designed to enroll 150 women, but accrual took longer than expected in part due to recurrent stock-outs of HIV testing supplies in Uganda. Reasons for ineligibility are shown in Figure 2.

**Figure 2.**
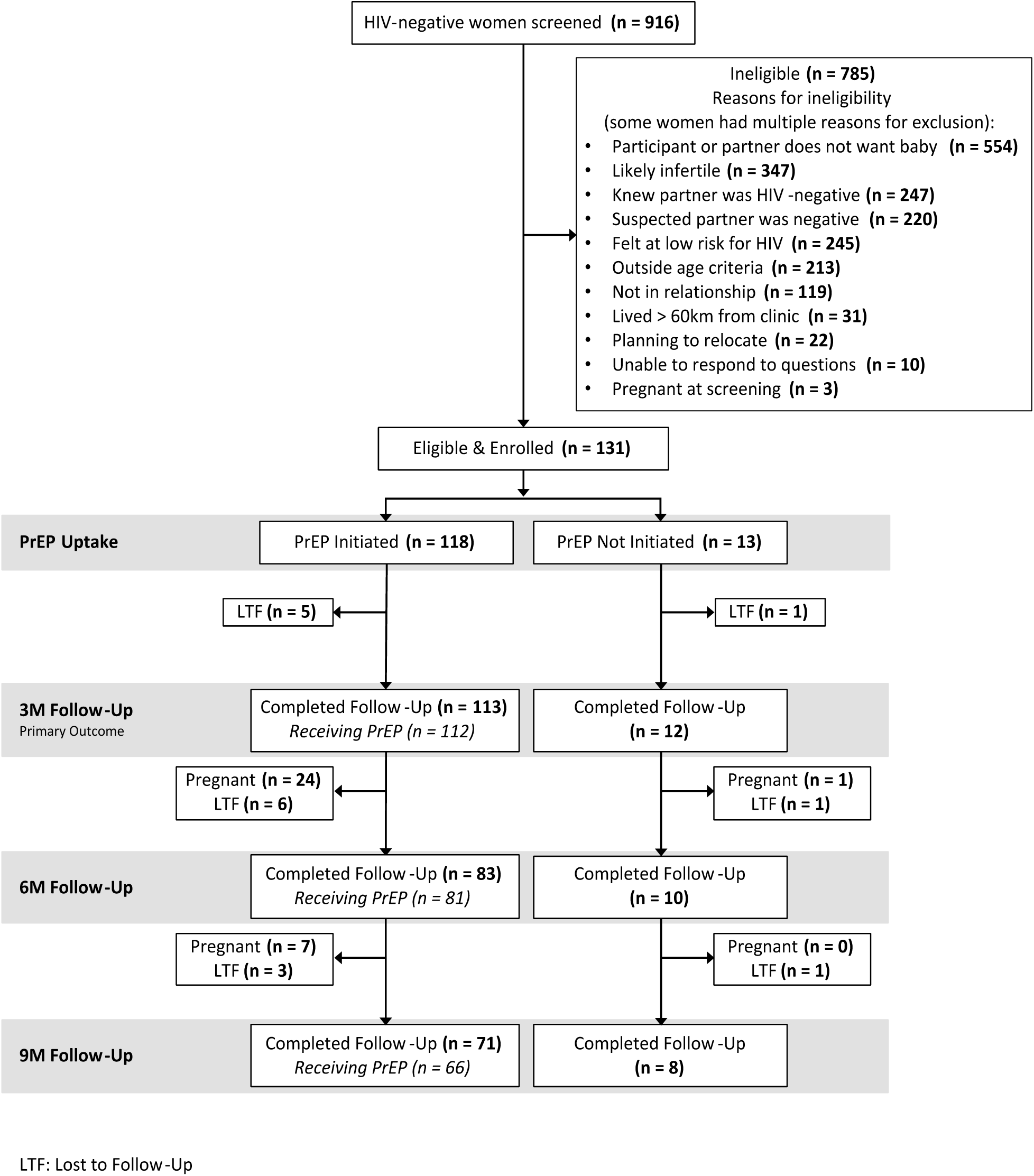
Summary of Screening, Eligibility, Enrollment, PrEP Uptake and Follow-Up

Table 1 presents the baseline characteristics of enrolled participants. Median (25^th^-75^th^ percentile) age was 28.7 (27.8-29.5) years with nearly all women (92%) reporting a prior pregnancy. Among women with a prior pregnancy, median (25^th^-75^th^ percentile) number of pregnancies was 2.9 (2.7-3.2). Most participants had completed primary school (68%), were currently employed (73%), married or living as married (94%), and reported that their partner or spouse was living with HIV (74%). Forty-five (35%) women screened positive for symptoms of depression, 14 (11%) reported problematic drinking within the past year, and perceived HIV risk score was high (21.3, 95% CI: 20.9-21.8).

**Table 1.**
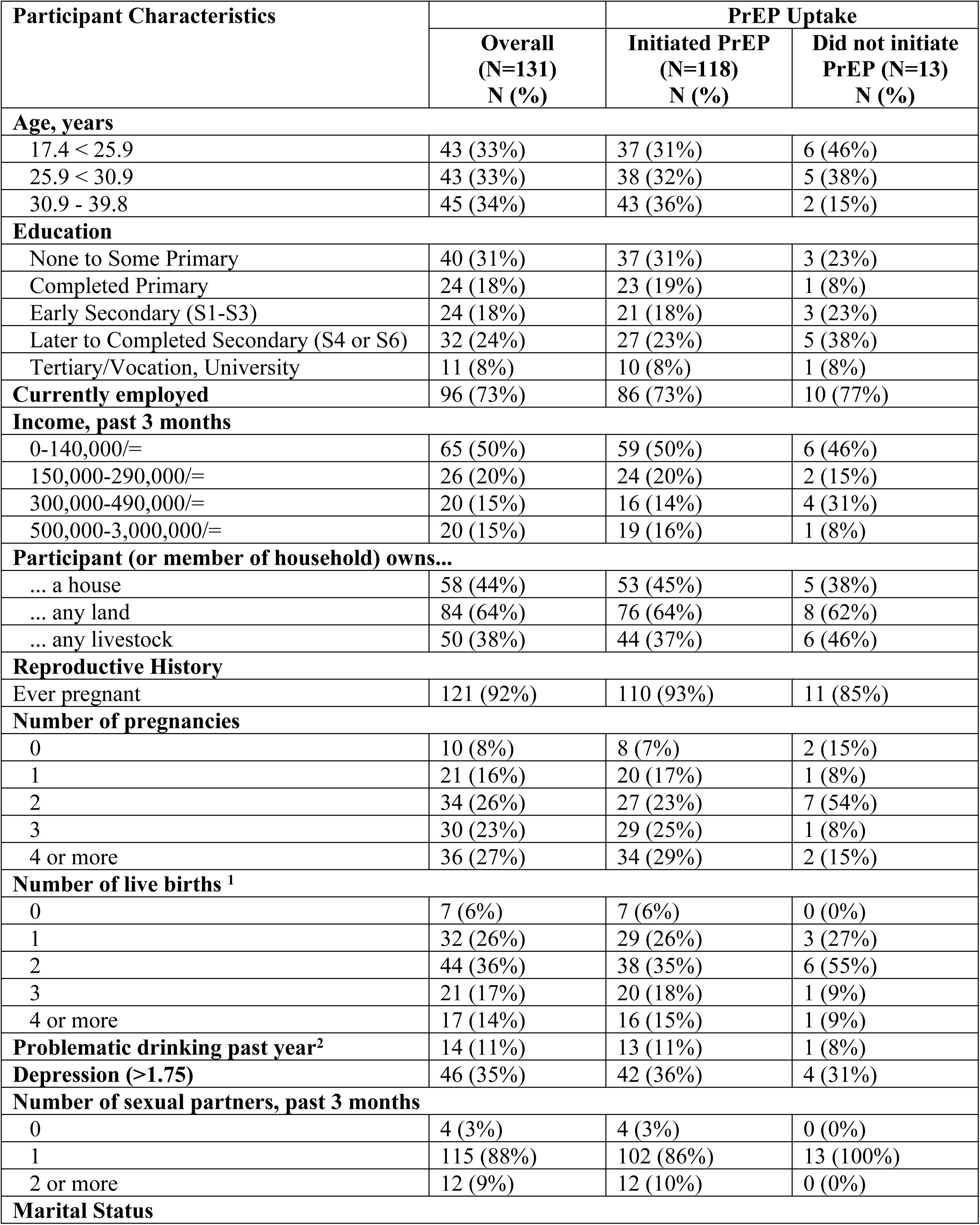
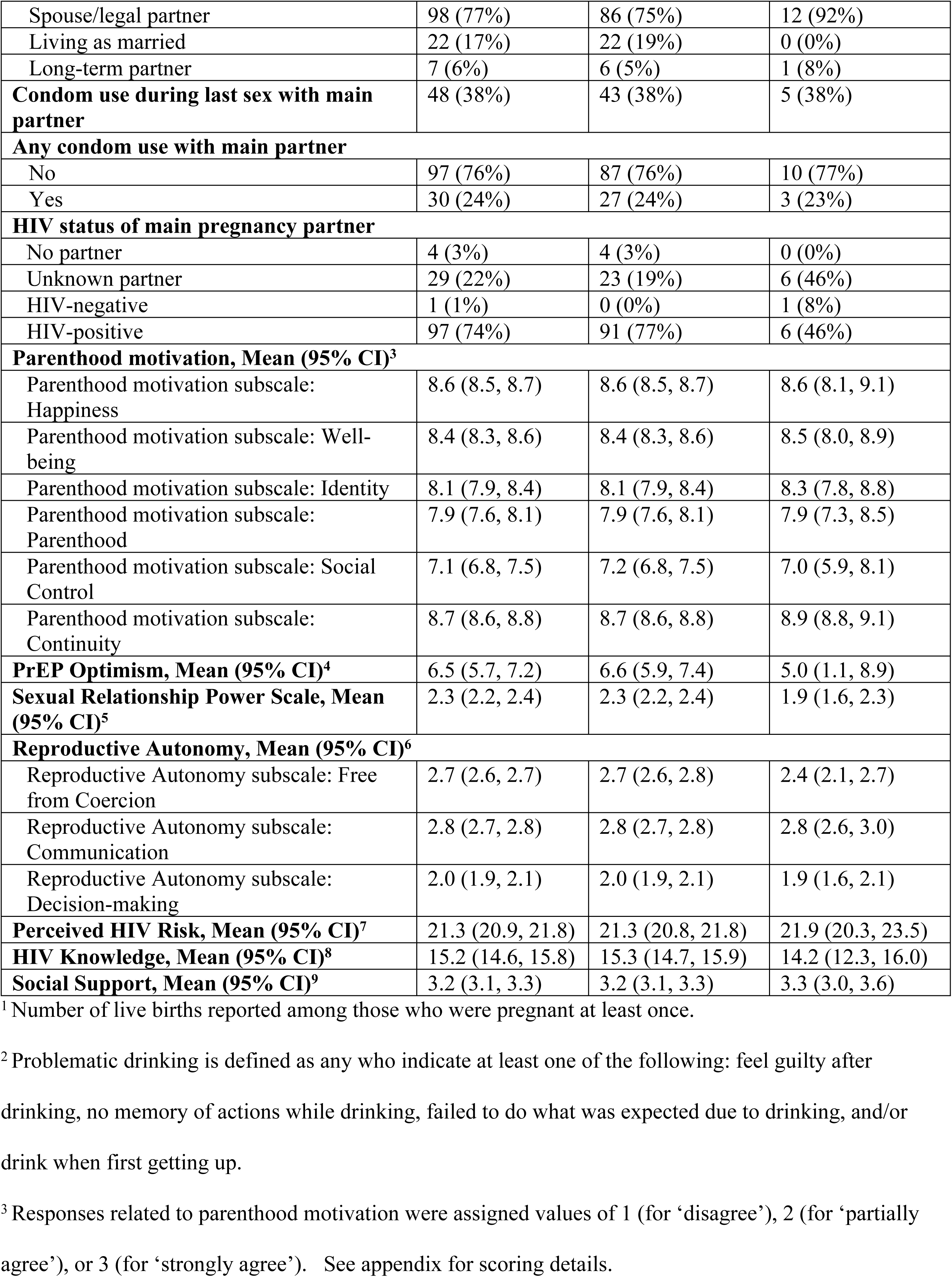
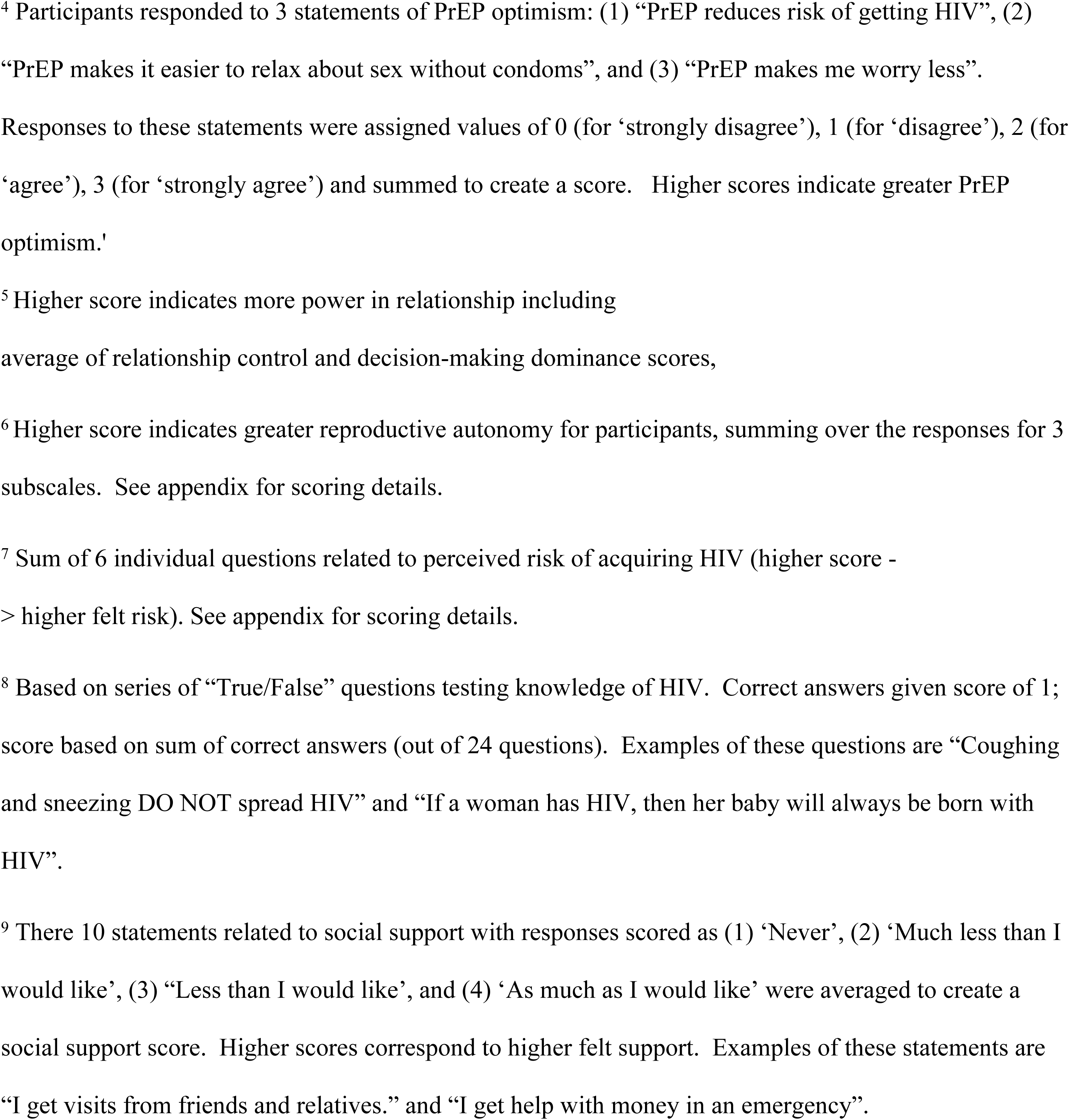
Baseline characteristics of N=131 participants in the Uganda Healthy Family Study, overall and according to PrEP uptake

**Table 2.** Univariable and multivariable-adjusted change in mean (95% confidence interval [CI]) adherence^a^ to PrEP during 3 months following initiation

| **Covariate** | **Univariable** |  | **Multivariable-adjusted<sup>b</sup>** |  |
| --- | --- | --- | --- | --- |
|  | **Change in percent adherence (95% CI)** | ***P*** | **Change in percent adherence (95% CI)** | ***P*** |
| Age per 5 years | 1.7% (-1.7%, 5.1%) | 0.33 | 1.7% (-1.7%, 5.1%) | 0.33 |
| Some secondary education or higher (vs none or primary education) | 2.4% (-4.6%, 9.4%) | 0.50 | 2.1% (-4.8%, 9.0%) | 0.55 |
| Number of live births (2+ vs 0,1) | 2.5% (-4.8%, 9.7%) | 0.50 | 1.0% (-6.9%, 8.9%) | 0.80 |
| Depression score (>1.75 vs ≤ 1.75) | -5.6% (-12.7%, 1.5%) | 0.12 | -5.6% (-12.7%, 1.5%) | 0.12 |
| Parenthood motivation subscale: Social Control | 0.04% (-1.8%, 1.9%) | 0.97 | 0.04% (-1.9%, 2.0%) | 0.97 |
| Sexual Relationship Power Scale | 1.1% (-4.2%, 6.5%) | 0.68 | 0.4% (-5.1%, 5.9%) | 0.89 |
| Reproductive Autonomy subscale: Decision-making | -2.6% (-9.5%, 4.3%) | 0.46 | -3.2% (-10.2%, 3.7%) | 0.36 |
| Perceived HIV Risk Score | -0.1% (-1.3%, 1.2%) | 0.90 | 0.04% (-1.2%, 1.3%) | 0.95 |

<sup>a</sup> Adherence measured as the percentage of time-stamped pillbox openings during active PrEP follow-up.<sup>b</sup> Multivariable-adjusted associations estimated using a covariate adjusted for confounders determined to be of interest using change-in-estimate methods considering all other covariates

Among 131 enrolled women, 17 (13%) moved or were otherwise lost to follow-up. A total of 53 pregnancies occurred among 131 women with 848 person-months of follow-up (median 8.8 months), resulting in a pregnancy incidence of 75% (95% CI: 57%, 98%) (Figure 2).

### PrEP Uptake

Of the 131 women enrolled, a total of 118 women (90%) initiated PrEP, all of whom chose to initiate at baseline. None had hepatitis B infection nor were ineligible based on serum creatinine. Those who initiated PrEP were older (mean age 29.0 vs. 25.7 years) and more likely to report having ≥3 lifetime pregnancies compared to those who did not initiate PrEP (54% vs. 23%). A greater proportion of women who initiated PrEP reported their partner was living with HIV compared to the proportion of women who did not initiate PrEP (76% vs. 46%). Mean perceived HIV risk score was lower among women who initiated PrEP compared to those who did not initiate PrEP (21.3 versus 21.9) and women who initiated PrEP in this study had higher PrEP optimistic beliefs compared to those that did not initiate PrEP (Table 1).

### Periconception PrEP Adherence

Among PrEP initiators, 101 (86% of N=118) had electronic adherence data through the first 3 months (Figure 2). Among these women, average adherence was 87% (95% CI: 83%, 90%) and 86 (85%) women had high adherence (≥80% of expected doses taken) through 3 months. PrEP adherence was not significantly associated with covariates of interest in adjusted models (Table 2, full model details in Appendix). Monthly adherence through 9 months was consistently high (Figure 3): in the first month average adherence was 86% (N=107; 95% CI: 83%, 90%) compared to the final month average adherence of 90% (N=48; 95% CI: 85%, 95%). Longitudinal models showed that month on study was not associated with adherence (data not shown). No women had provider-directed PrEP holds or stops for renal dysfunction or other clinical events.

### Periconception Drug Concentrations

TFV concentrations were processed from 112 participants contributing 44 plasma and 104 dried blood spot samples at 3 months. At 6 months, 25 plasma and 79 dried blood spot samples collected from 81participants were processed. At 9 months, 22 plasma and 65 dried blood spot samples collected from 66 participants were processed. All processed samples were from women who were not pregnant at the time of collection.

Plasma TFV ≥40ng/mL was observed in 66%, 56%, and 45% of samples collected at 3, 6, and 9 months, respectively (Figure 4). Electronic adherence data from 3 and 30 days prior to sample collection were significantly correlated with plasma TFV levels at the 3-month visit (*ρ=*0.45 *P*=0.006 for 3 days and *ρ=*0.44 *P*=0.01 for 30 days). Correlations at 6 and 9 months were also moderate to high and statistically significant (data not shown), suggesting that pill-taking behavior over longer time frames correlates well with objective adherence behavior reflecting pill taking behavior over the past week. TFV-DP (indicative of use during past 6-8 weeks) concentrations of ≥600 fmol/punch were detected among 47% of 3-month, 41% of 6-month, and 45% of 9-month samples (Figure 4). Electronic adherence data and 3-month TFV-DP levels did not correlate with pillcap data collected during the 30 (*ρ*=0.06; *P*=0.55) and 60 (*ρ=*0.17; *P*=0.11) days prior to sample collection. Correlations at 6 and 9 months were similarly low and not statistically significant (data not shown). Figure 4 presents the proportion of participants with high, medium, low, and undetected adherence by pillcap, plasma, and dried blood spot at 3, 6 and 9 months of follow-up.

**Figure 3a.**
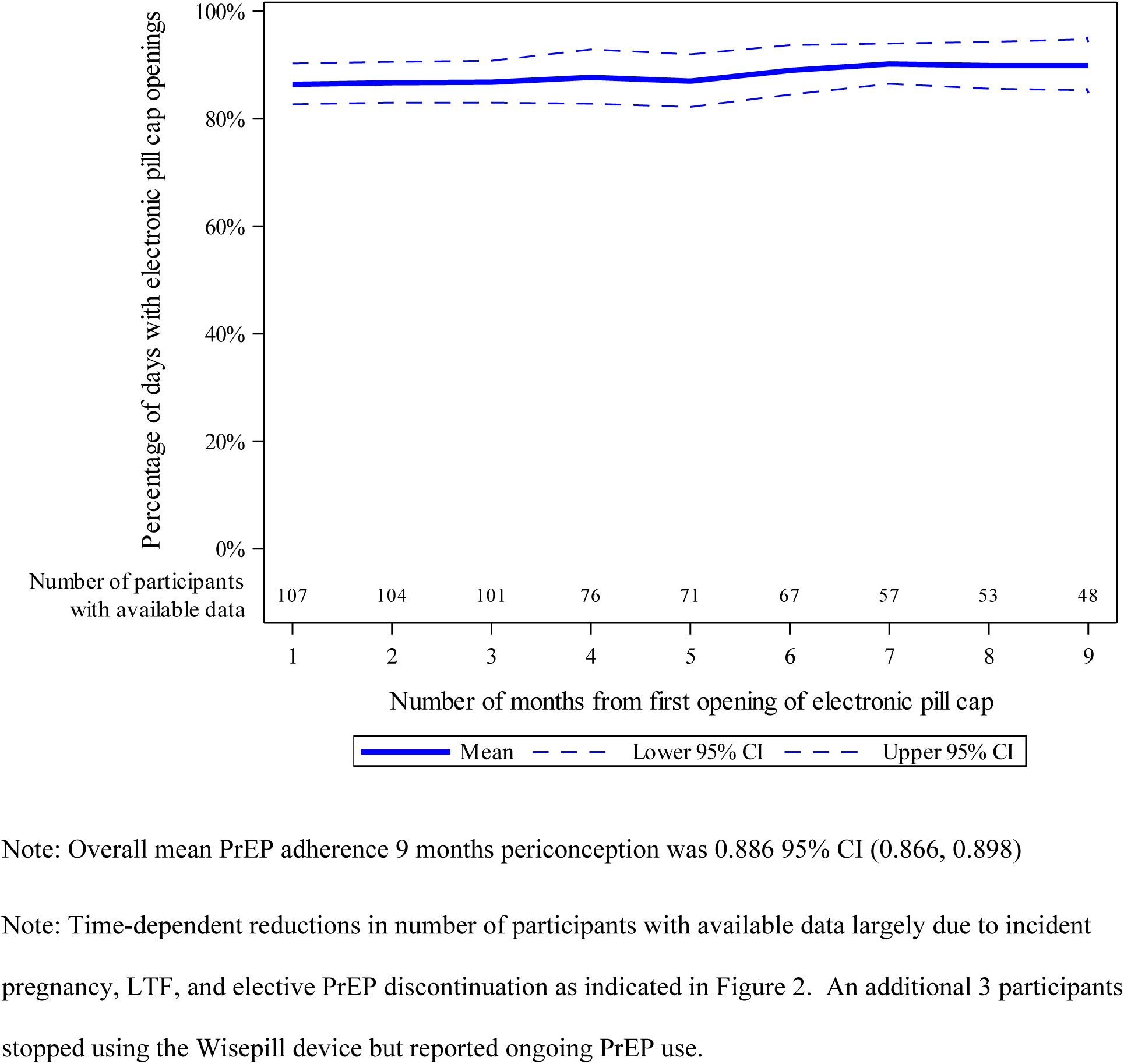
Mean and 95% confidence interval bands for electronic adherence to PrEP during periconception period over time.

**Figure 3b.**
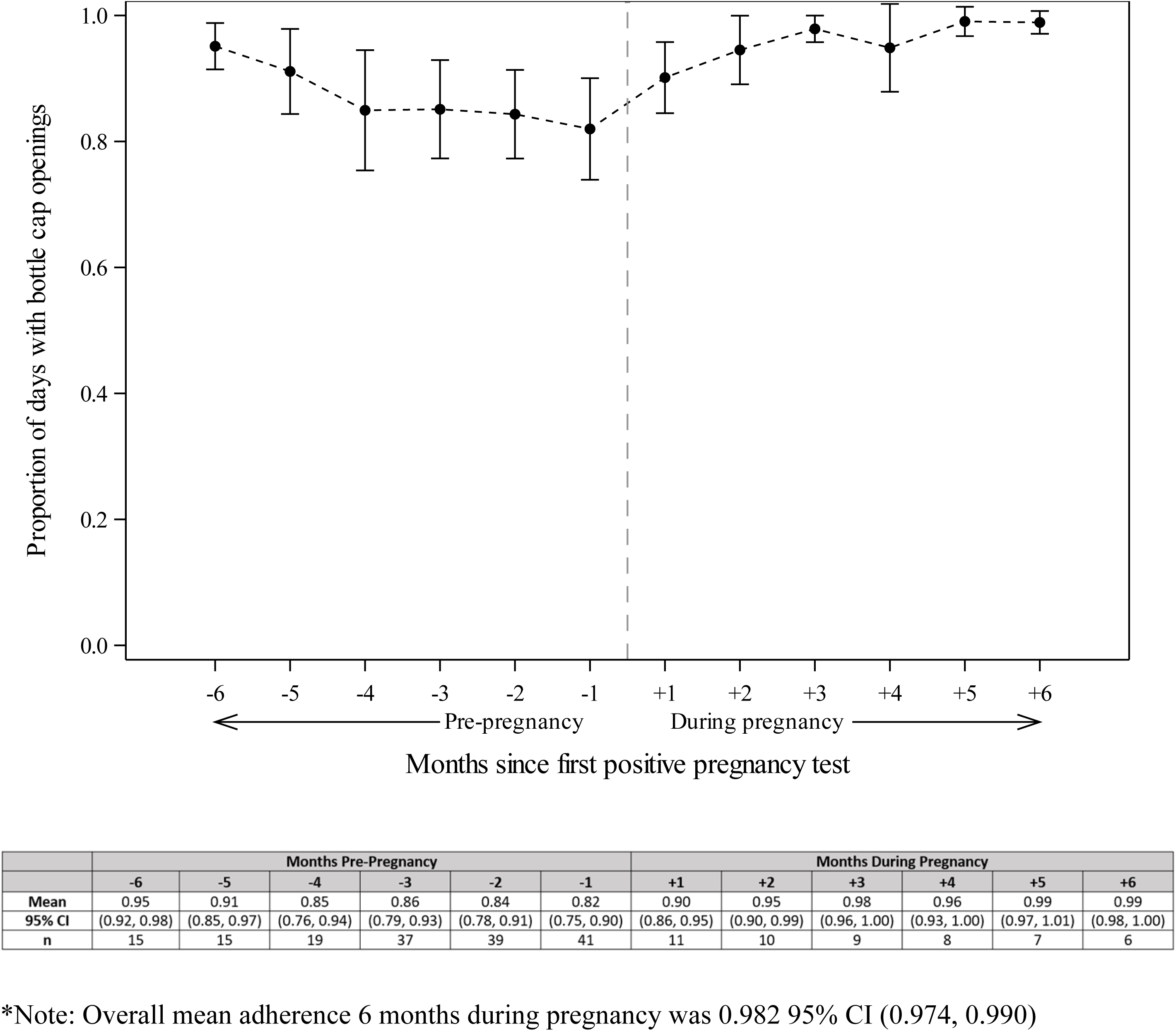
Adherence prior to and post-pregnancy among those women with incident pregnancy

**Figure 4.**
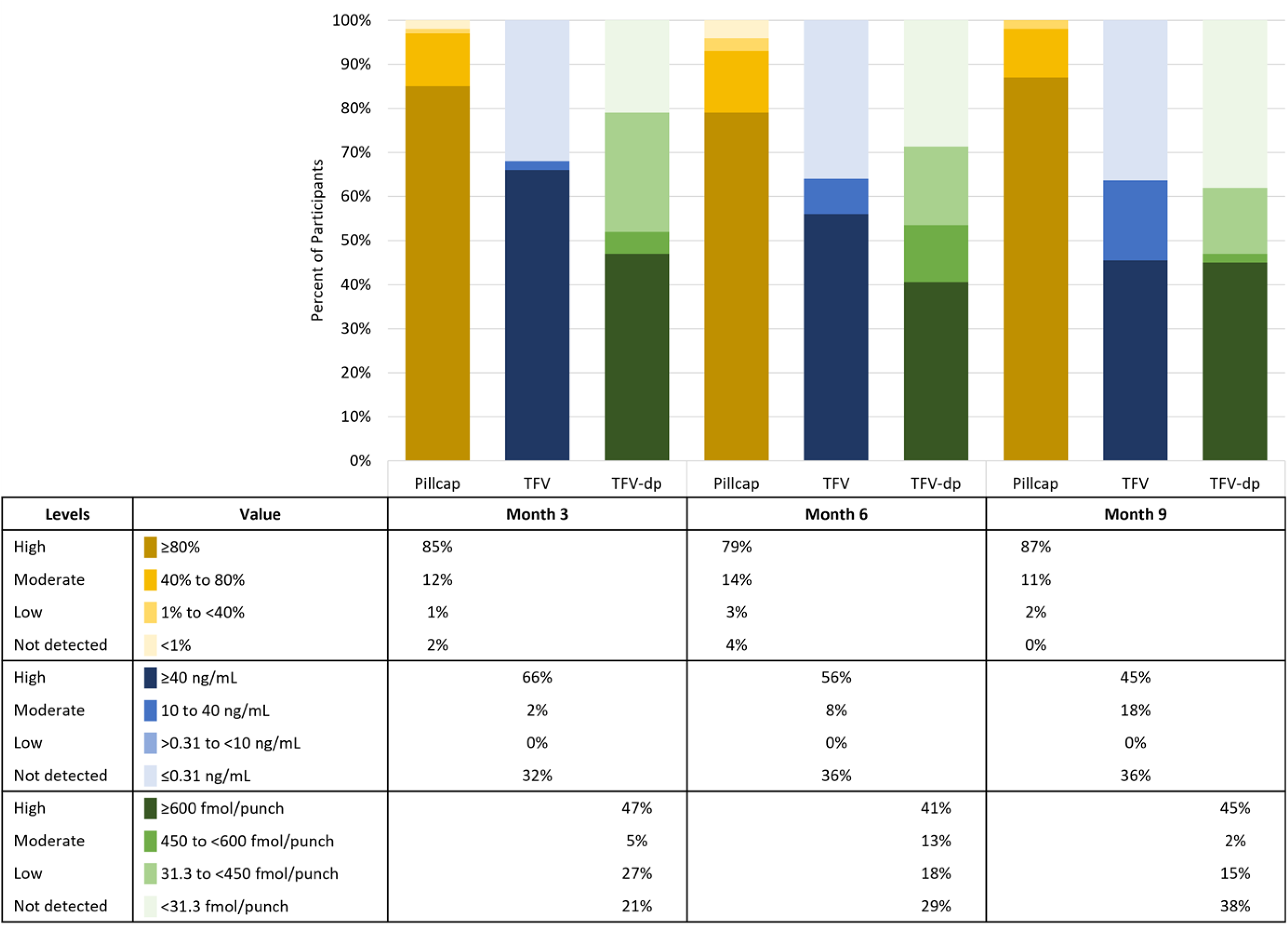
Periconception adherence at 3, 6, and 9 months using 3 assessment methods.

### HIV Incidence

One participant tested positive for HIV at 9 months. This participant was not pregnant. Although electronic adherence data suggested high PrEP adherence (>80% of doses all months), plasma TFV levels were undetectable at 3, 6, and 9 months of follow-up.

### Pregnancy PrEP Use

Forty-eight (91%) of 53 pregnant women ever used PrEP; of whom 35 (66%) completed exit procedures at the incident pregnancy visit, 17 (32%) were followed through pregnancy outcome.. Overall mean adherence by electronic pillcap during the first 6 months of pregnancy was 98% (95% CI: 97%, 99%). Figure 3b highlights consistent adherence across periconception and pregnancy time periods. Of the 17 pregnancies with outcome information, 12 (71%) resulted in live births and 5 (29%) resulted in miscarriages or stillbirth or termination.

## Discussion

We observed high PrEP uptake and use among periconception and pregnant women in Uganda participating in the Healthy Families-PrEP counseling intervention. These are the first data we are aware of showing high uptake and high, sustained adherence (by pillcap and plasma TFV) to daily, oral TDF/FTC as PrEP in a population of women with indications for HIV prevention in Uganda. Over 90% of participants chose PrEP as part of a safer conception strategy and mean pillcap adherence over 3 months was 87% (95% CI: 83%, 90%). In addition, electronic adherence demonstrated persistent adherence behavior over 9 months of periconception as well as pregnancy follow-up. Despite high pregnancy (35%) and STI (21%) incidence at 6 months (46), only a single HIV infection occurred (out of >70 person-years of follow-up). Our short term (plasma TFV) and longer term (intra-erythrocytic TFV-DP) data suggest that closer to half of women consistently took oral PrEP. This proportion is high compared to many other implementation studies of PrEP for women, as outlined in the introduction, and higher than to the TDF/FTC arm of the recent HPTN-084 trial in which 42% of women had high plasma TFV and 18% had high TFV-DP levels above 700fmol/punch (65); these findings highlight opportunities of PrEP for periconception and pregnancy prevention. Differences observed across 3 objective measures of PrEP adherence in this cohort emphasize the value of incorporating multiple adherence measurements that capture different time exposures while revealing gaps in our understanding of tenofovir drug-level data interpretation for cis-gender African women who are neither pregnant nor post-partum.

Emerging studies of PrEP use among adolescent girls, and young women have mixed results, although most note high PrEP initiation and declining use over time (66–70). The high uptake and sustained adherence observed in this study are consistent with a recent periconception study conducted among HIV-negative Kenyan women within a serodifferent partnership and desire to have a child (27). In the Kenyan pilot study, PrEP was initiated by 100% of the HIV-negative partners (including 40 couples in which the woman was HIV-negative) and 81% took at least 80% of PrEP doses one month prior to pregnancy based on electronic pill caps (27). Another recent periconception study in the U.S. observed that among 25 women taking PrEP while seeking conception with a male partner living with HIV, 87% had TFV-DP levels consistent with taking at least 4 doses/week (71). Indeed, our qualitative sub-study with participants and partners suggested that the motivation to have a safe option to fulfill the cultural expectations of having children while maintaining sero-different relationships encouraged PrEP adherence (72). Conversely, a prospective cohort study in South Africa to assess the uptake and effectiveness of a safer conception intervention found lower PrEP initiation at 51% (22 of 43) among HIV-negative women in sero-different or unknown serostatus relationships, but this study was conducted early in South Africa’s PrEP roll-out (73). Overall our data and the literature suggest that the periconception time period, when women may be motivated to achieve reproductive goals and deliver an HIV-uninfected baby may help them to overcome oral PrEP adherence challenges (34).

More data have been collected from women using PrEP during pregnancy with conflicting results (31, 32, 74). In an implementation project in Kenya, 22% of 9,376 pregnant and postpartum women and 79% of 193 pregnant and postpartum women with a partner known to be living with HIV initiated PrEP. Only 39% of these women continued to use PrEP after the first month (31). In South Africa, in a cohort of 1201 HIV-negative, pregnant women accessing care at a public sector clinic, 84% chose to use PrEP and 58% returned for 3-month follow-up (74). Among those women, just 19% of pregnant women and 11% of postpartum women had drug concentrations consistent with taking 2-6 doses per week (75). We observed ongoing high adherence, by pillcap, among women during pregnancy, suggesting that starting PrEP prior to pregnancy may provide an opportunity to overcome adherence barriers prior to onset of pregnancy symptoms or simply a selection bias that women who choose to use periconception PrEP are likely to have ongoing success. While the periconception period is not routinely identified in general clinical care, we maintain that this may be a key opportunity to engage women in HIV prevention, including PrEP care.

The Healthy Families PrEP intervention offered counselling delivered by local lay counselors to support daily PrEP use in the context of planning for or being pregnant (Figure 1). Quarterly adherence support was adapted from the Lifesteps intervention, and utilized education, problem solving and motivational interviewing strategies to promote adherence. Lifesteps was first adapted for PrEP among HIV-serodifferent couples in Uganda and successfully promoted PrEP adherence (45). While this counseling approach was effective for most women in our cohort, some encountered ongoing adherence challenges. Future iterations of this work should aim to predict who which women are most likely to experience adherence challenges, and identify what role adherence counseling may play in supporting prevention effective PrEP use, regardless of PrEP formulation (e.g. adaptive interventions (79)).

Interpretation of our data varies to some extent by the adherence measure used and the degree to which a given adherence threshold correlates with PrEP effectiveness. We present various approaches to data interpretation to learn from the data, while recognizing these limitations. We categorized ≥80% pillcap adherence as high based on trends in the literature and associations with protection (58, 59). *In vitro* pharmacokinetic (PK) data suggest that 6-7 doses per week may be required to achieve stable FTC and TFV drug concentrations in cervical and vaginal mucosal tissues (86-100% of doses); however, which compartment requires what drug concentration to achieve protection remains unclear for HIV via receptive vaginal intercourse(59). We categorized concentrations of ≥40ng/mL for plasma TFV as high informed by data that includes African women, suggesting protection from this level, which reflects dosing in the past 24 hours (20, 60). Whole blood concentrations of TFV-DP associated with protection from HIV for women are not available. Twenty women from East and Southern Africa who were 6-12 weeks post-partum and taking directly-observed oral PrEP 7 times per week had 25^th^ percentile concentrations of TFV-DP of 1053 fmol/punch (63), and the corresponding estimate for 4 doses per week was 600fmol/punch. For 25 U.S. women (5 of whom were Black, none noted to be pregnant), the 25^th^ percentile concentration for steady state dosing at 4 doses per week was approximately 700 fmol/punch (80); these data, plus data from men, has informed use of a concentration of 700 fmol/punch to indicate high adherence (63, 80–82). However, because data suggest that this cut-off is not sensitive for adherence in African cis-gender women (83), we used 600 fmol/punch.

Differences observed across the PrEP adherence measures in this cohort highlight the value of incorporating multiple adherence measurements and expose some gaps in our understandings of tenofovir drug-level data interpretation for cis-gender women. Importantly, adherence by any one of these measures is higher than seen in many prior cohorts of women of reproductive age. Further, one HIV seroconversion was observed despite high pregnancy and STI incidence indicating that the intervention supported women to avoid HIV transmission over time(84). Exploring possible reasons for discrepancies across measures is important for interpreting the effect of our intervention and informing future analyses. First, the categories of high are not equivalent across the measures in terms of dosing or time period: 80% of pills taken by electronic pillcap equates to 5-6 doses per week averaged over ∼90 days, ≥40ng/mL represents dosing in the last 24 hours, and ≥600 fmol/punch TFV-DP represents 4 or more doses per week over 6-8 weeks. In addition, the samples represent different groups: pillcap data were available for 83%, plasma data were processed for 39%, and whole blood samples were processed for 93% of women accessing PrEP at the 3-month (primary outcome) timepoint. High adherence by electronic device persisted whereas the proportion with high levels waned over time with plasma and whole blood measures. Women may have persisted using the pillcap device without taking pills to please the researchers or counselors (social desirability bias)(85). However, studies have shown that adhering to pill cap opening without taking medication is difficult to maintain over time(86); further, PrEP use was not required to remain in the study. Plasma TFV measures pill-taking in the past week and women may have been prompted to take doses by reminders about upcoming clinic visits thus explaining higher plasma levels even when TFV-DP concentrations were lower; however, correlations between plasma TFV and electronic pillcap data suggest that pillcap use over 30 days aligned with an objective, biologic measure in the week prior to blood draw. Correlations between electronic pillcap and whole blood (DBS) were low. In a secondary analysis of whole blood spots collected from men and women with electronic pillcap data in the Partners PrEP demonstration project, TFV-DP measurements were specific but not sensitive for electronic pillcap adherence (83) with sensitivity lowest for the highest categories of MEMS adherence.

Those authors speculate that the discrepancy may relate to interindividual differences in drug disposition (87), (88, 89). While inter-individual differences in PK are certain, intra-individual PK changes over time are less likely (particularly with respect to anemia, BMI and other biological factors impacting drug disposition.) Thus the downward trends of DBS TFV-DP and plasma TFV values likely at least partially reflect behavioral fatigue with daily pill taking and highlight the importance of adherence support for daily PrEP use. The most conservative estimates of adherence also suggest that some women may require different adherence support than what was provided in this study and ongoing work is needed to optimize adherence support for women (79, 90). Again, adherence by all measures was higher in this cohort than for most PrEP implementation cohorts among women.

We did not identify factors that predicted PrEP use over time in our models. Covariates were selected based on our conceptual framework and further refined by our qualitative data and the literature of factors associated with PrEP use in other cohorts of women. We may have been under-powered to detect associations, particularly because pillcap adherence was consistently high for most women (confidence intervals for associations were wide.). In terms of uptake, we observed that women who initiated PrEP in this study had higher PrEP optimistic beliefs compared to those that did not initiate PrEP (based on average PrEP optimism score). Our qualitative data also suggested a high perception of PrEP effectiveness among participants (72). This finding aligns with a recent study conducted in Central Uganda showing high acceptability and willingness to take PrEP if offered among high-risk populations in Uganda (91). In their study, which included a two-day training workshop on PrEP for health care workers, investigators observed an increase in PrEP interest and knowledge 9 months post intervention.

In many trials of PrEP for women, male partner engagement is associated with higher adherence (92–95). Due to the challenges with HIV-serostatus disclosure globally and the desire to offer HIV prevention to women who may not know her partner’s serostatus or be able to engage him in HIV prevention, our study was designed to enroll women as individuals. The absence of required partner involvement does not appear to have dampened enthusiasm for PrEP use. From our qualitative data, some participants reported that the safer conception program encouraged them to disclose their serostatus with partners, support each other to ensure daily medication adherence, and offered a sense of hope to *“fight”* the virus together (96). Given the limited prevention options for women who choose to conceive with men with HIV, WHO guidelines identify serodifferent couples considering conception as a priority group for PrEP (97). The FDA labeling information and the U.S. Perinatal ART Guidelines support periconception PrEP use (34, 98–102). Similarly, WHO and CDC guidelines emphasize that eliminating perinatal transmission requires pre-conception counseling to reduce transmission to the mother and therefore the child (102, 103).

Strengths of this study include the unique population, use of prospective data, use of well-validated tools for evaluating potential social and behavioral factors that could influence PrEP use, and objective measurements of PrEP adherence using daily electronic monitoring device and plasma and whole blood drug levels. However, this study also has limitations. First, due to intermittent country-wide stock-outs of HIV testing kits, attendance at HIV testing centers was low during much of the recruitment period, resulting in slower than expected recruitment. Second, while PrEP was being rolled out in this clinic in Uganda at the time of our study, women could access enhanced counseling and personalized pharmacy services in our program. Therefore, women may have enrolled in our program regardless of pregnancy plans but to access PrEP outside of the ART pharmacy programs where stigma may impair uptake. Finally, due to funding restrictions and study design, we have limited pregnancy data follow-up.

In conclusion, PrEP offers a desirable and effective tool for HIV prevention among HIV-negative women who could acquire HIV during periconception and pregnancy periods. These data suggest that women planning for and with pregnancy should be prioritized for PrEP implementation and adherence support, particularly in settings with high fertility rates and generalized HIV epidemics. Future work will aim to evaluate implementation of the Healthy Families PrEP intervention on periconception, pregnancy, and post-partum PrEP use in public sector clinics in Uganda.

## Data Availability

The data will be available in the Harvard Dataverse database after acceptance for publication.

## Acknowledgements

We gratefully acknowledge the contributions of the participants, families, and the entire study team for their contributions. This work is dedicated to the memory of our dear colleague, mentor, and friend, Dr. Mwebesa Bosco Bwana, who led our Healthy Families team and this study with deep compassion for the patients he loved to serve.

## Appendix A: Table 1 Scoring Details

### Parenthood Motivation

A score was derived by summing over 3 statements related to each subscale as follows:

• ***Happiness***: (1) “It is nice to have children around” + (2) “I want to have a unique relationship with the child”’ + (3) “Bring up children brings happiness”
• ***Well-being***: (1) “Parenthood makes the relationship with your partner complete” + (2) “Children make life complete” + (3) “Parenthood gives you a goal to live for
• ***Identity***: (1) “It is obvious to have children” + (2) “Parenthood is a sign of being grown up” + (3) “Parenthood is the nature of women”
• ***Parenthood***: (1) “Parenthood fulfills motherly feelings” + (2) “Parenthood is satisfying” + (3) “I want to experience pregnancy and birth”
• ***Social Control***: (1) “My environment (others, family) expects it of me” + (2) “Others around me have children” + (3) “I want to have a baby to avoid being an outsider”
• ***Continuity***: (1) “Parenthood allows a person to continue the family name/tradition” + (2) “Parenthood allows a person not to be alone when you are old” + (3) “I want to have something of myself that continues living after I die”

### Reproductive Autonomy

• ***Free from Coercion*** responses to the following 5 statements were scored as (1) for ‘strongly agree’, (2) for ‘agree’, (3) for ‘disagree’, (4) for ‘strongly disagree’: (1) “My pregnancy partner has stopped me from using a method to prevent pregnancy when I wanted to use one”, (2) “My pregnancy partner has messed with or made it difficult to use a method to prevent pregnancy.”, (3) “My pregnancy partner has made me use a method to prevent pregnancy when I did not want to use one.”, (4) “If I wanted to use a method to prevent pregnancy, my desired pregnancy partner would stop me.”, and (5) “My pregnancy partner has pressured me to become pregnant.”
• ***Communication*** responses to the following 5 statements were scored as (1) for ‘strongly disagree’, (2) for ‘disagree’, (3) for ‘agree’, (4) for ‘strongly agree’: (1) “My pregnancy partner would support me if I wanted to use a method to prevent pregnancy.”, (2) “It is easy to talk about sex with my pregnancy partner.”, (3) “If I didn’t want to have sex with I could tell my pregnancy partner.”, (4) “If I was worried about being pregnant or not being pregnant, I could talk to my pregnancy partner about it.”, (5) “If I really did not want to become pregnant, I could get my pregnancy partner to agree with me.”
• ***Decision making*** responses to the following 4 statements were scores as (1) for ‘My partner or someone else’, (2) for ‘Me and my pregnancy partner (or someone else) equally’, (3) for ‘Me’: (1) “Who has the most say about whether you use a method to prevent pregnancy?”, (2) “Who has the most say about which method you would use to prevent pregnancy?”, (3) “Who as the most say about when you have a baby in your life?”, and (4) “If you become pregnant but it was unplanned, who would have the most say about whether you would raise the child, seek adoptive parents, or have an abortion?”

### Perceived HIV Risk

• (1) “What is your gut feeling about how likely you are to get infected with HIV?” *[responses range from (1) Extremely unlikely to (4) Extremely likely]*
• (2) “I worry about getting infected with HIV” *[responses range from (1) Never to (4) All the time]*
• (3) “Getting HIV is something I am….” *[responses range from ‘Not concerned about’ to ‘Extremely concerned about’]*
• (4) “I am sure I will not get infected with HIV.” *[responses range from (1) ‘Strongly agree’ to (4) ‘Strongly disagree’]*
• (5) “I feel I am unlikely to get infected with HIV.” *[responses range from (0) Strongly agree through (5) Strongly disagree]*
• (6) “I feel vulnerable to HIV infection.” *[responses range from (5) Strongly agree through (0) Strongly disagree]*

## Appendix B: Full Models for Table 2

**Appendix Table 1.**
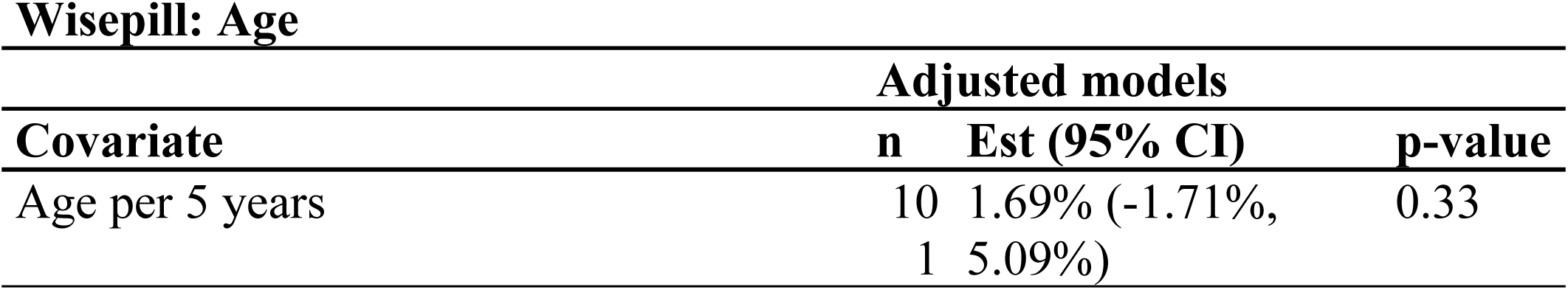
Adherence through 91 days measured through Wisepill: Age.

**Appendix Table 2.**
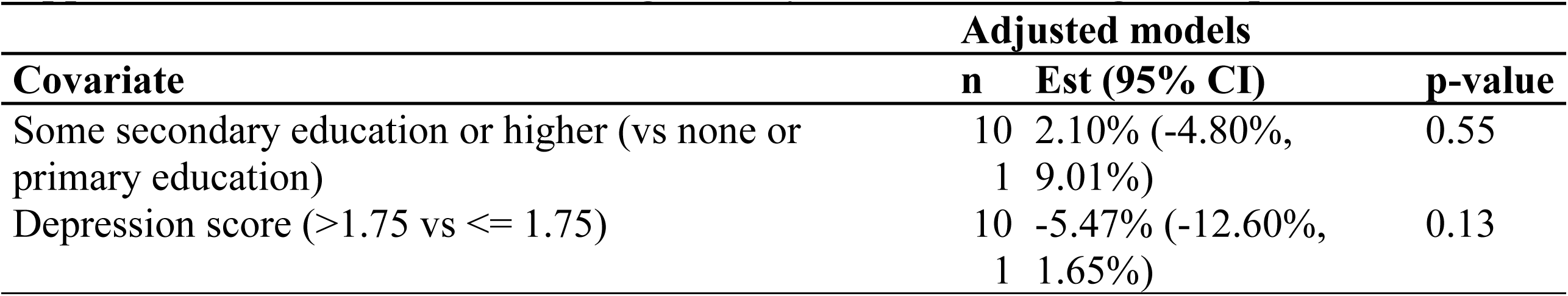
Adherence through 91 days measured through Wisepill: Education.

**Appendix Table 3.**
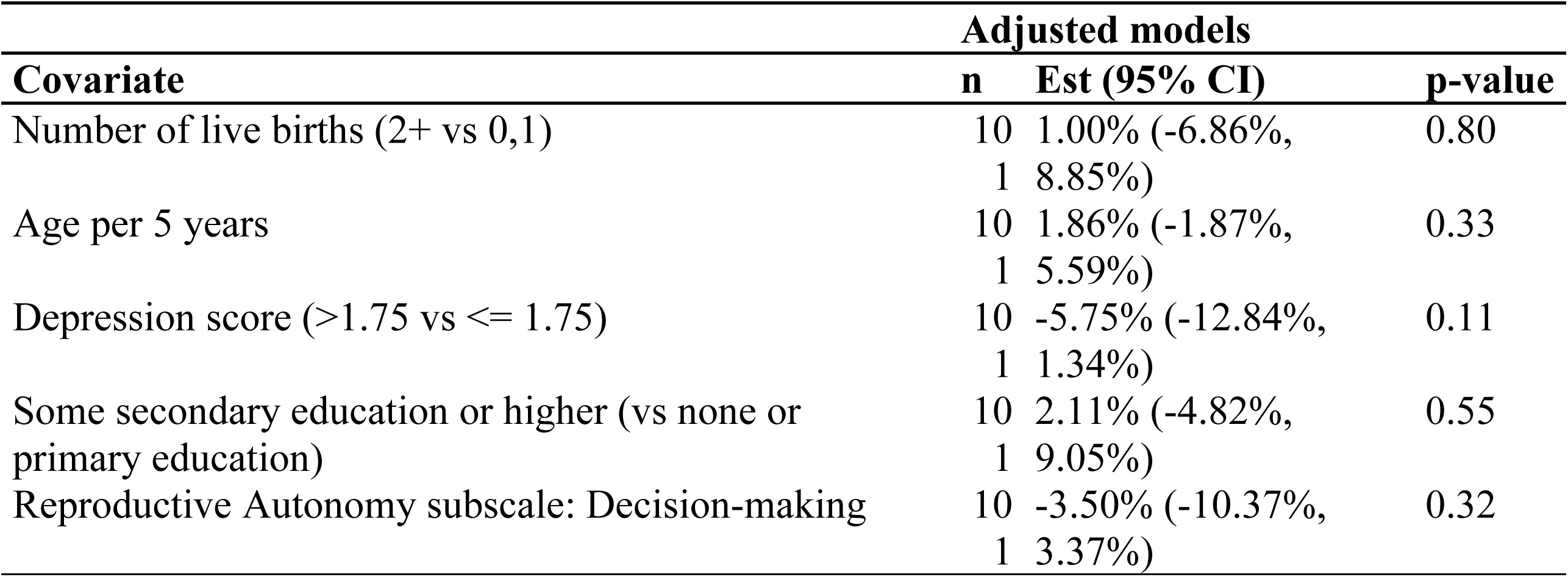
Adherence through 91 days measured through Wisepill: Number of live births.

**Appendix Table 4.**
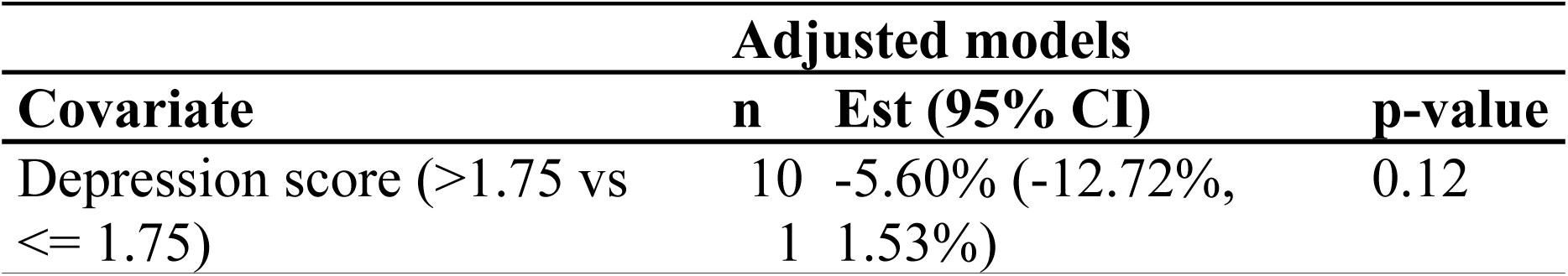
Adherence through 91 days measured through Wisepill: Depression.

**Appendix Table 5.**
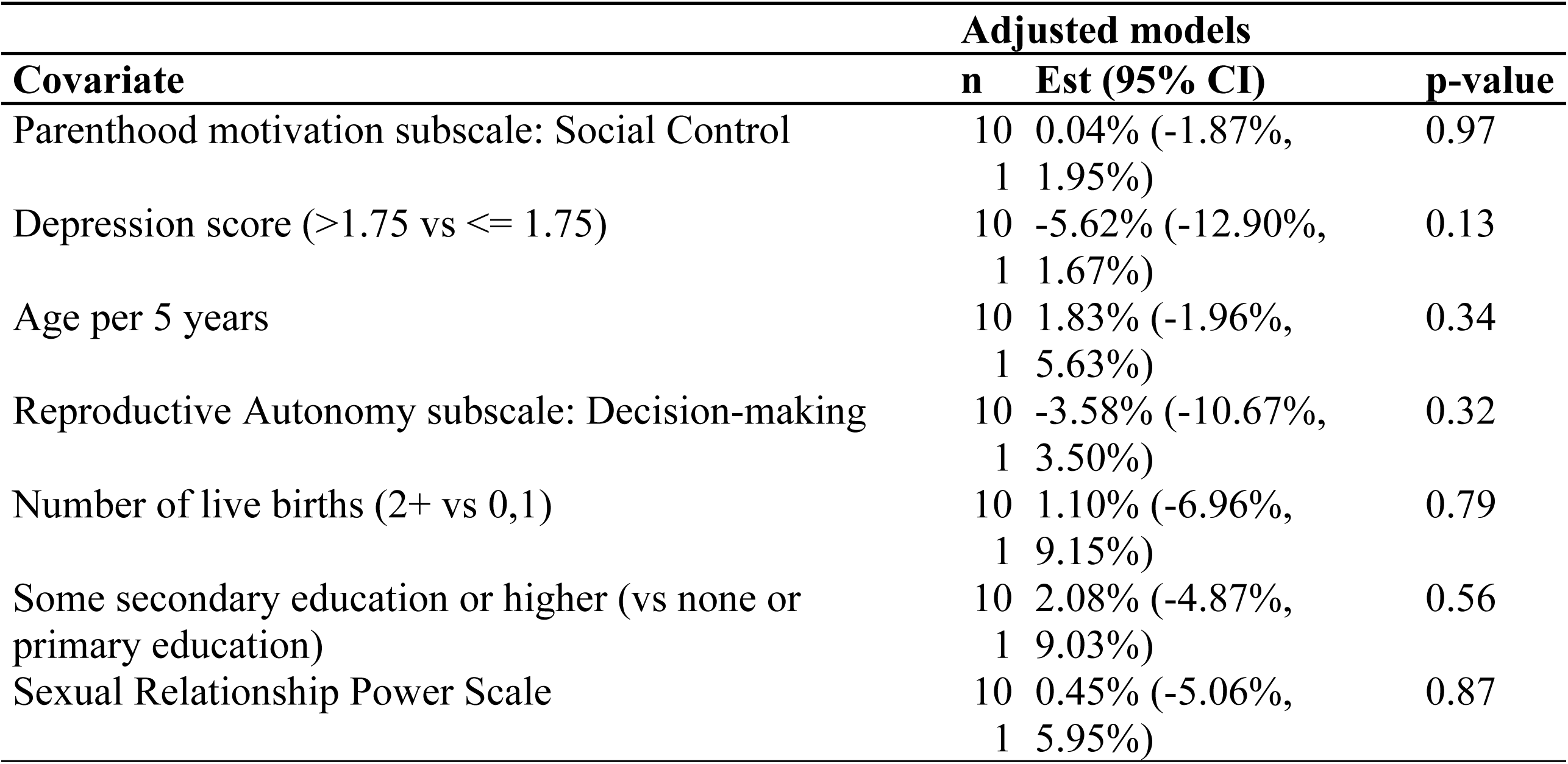
Adherence through 91 days measured through Wisepill: Parenthood motivation subscale: Social Control.

**Appendix Table 6.**
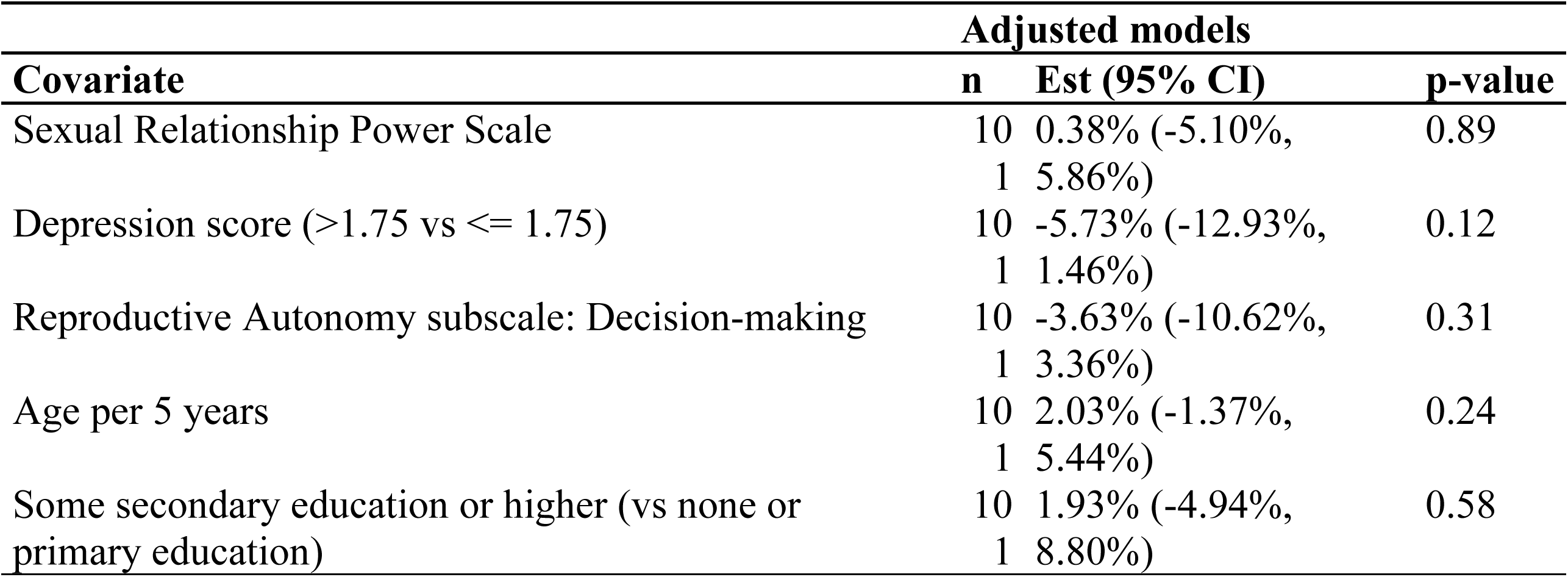
Adherence through 91 days measured through Wisepill: Sexual Relationship Power Scale.

**Appendix Table 7.** Adherence through 91 days measured through Wisepill: Reproductive Autonomy subscale: Decision-making.

| **Covariate** | **Adjusted models** |  |  |
| --- | --- | --- | --- |
|  | **n** | **Est (95% CI)** | **p-value** |
| Reproductive Autonomy subscale: Decision-making | 101 | -3.23% (-10.19%, 3.73%) | 0.36 |
| Age per 5 years | 101 | 1.95% (-1.48%, 5.38%) | 0.26 |

**Appendix Table 8.** Adherence through 91 days measured through Wisepill: Perceived HIV Risk Score.

| **Covariate** | **Adjusted models** |  |  |
| --- | --- | --- | --- |
|  | **n** | **Est (95% CI)** | **p-value** |
| Perceived HIV Risk Score | 10<br>1 | 0.04% (-1.22%,<br>1.30%) | 0.95 |
| Age per 5 years | 10<br>1 | 1.88% (-1.89%,<br>5.66%) | 0.33 |
| Depression score (>1.75 vs ≤ 1.75) | 10<br>1 | -5.77% (-12.87%,<br>1.34%) | 0.11 |
| Reproductive Autonomy subscale: Decision-making | 10<br>1 | -3.48% (-10.37%,<br>3.41%) | 0.32 |
| Some secondary education or higher (vs none or primary education) | 10<br>1 | 2.13% (-4.83%,<br>9.10%) | 0.55 |
| Number of live births (2+ vs 0,1) | 10<br>1 | 0.96% (-6.96%,<br>8.89%) | 0.81 |

